# Immune response to third SARS-CoV-2 vaccination in seronegative kidney transplant recipients: possible improvement by mycophenolate mofetil reduction

**DOI:** 10.1101/2022.01.18.22269420

**Authors:** Marta Kantauskaite, Lisa Müller, Jonas Hillebrandt, Joshua Lamberti, Svenja Fischer, Thilo Kolb, Katrin Ivens, Michael Koch, Marcel Andree, Nadine Lübke, Michael Schmitz, Tom Luedde, Hans Martin Orth, Torsten Feldt, Heiner Schaal, Ortwin Adams, Claudia Schmidt, Margarethe Kittel, Eva Königshausen, Lars C. Rump, Jörg Timm, Johannes Stegbauer

## Abstract

**Background:** Modification of vaccination strategies is needed to improve the immune response to SARS-CoV-2 vaccination in kidney transplant recipients (KTRs).

**Methods:** This multicenter observational study aimed to determine antibody kinetics among 60 seropositive KTRs and analyzed the effects of the third vaccination against SARS-CoV-2 in 174 previously seronegative KTRs. We investigated whether mycophenolate mofetil (MMF) dose reduction by 25-50% prior the third vaccination influences vaccination success.

**Results:** 18 of 60 (30%) seropositive KTRs became seronegative in the serological assay within six months. Loss of antibodies was predicted by low initial antibody levels (≤206.8 BAU/ml), older age, and impaired graft function. A third vaccination in previously seronegative KTRs induced seroconversion in 56 of 174 (32.1%) KTRs with median antibody levels 119 (76–353) BAU/ml and median neutralizing capacity titer of 1:10 (0– 1:40). Multivariate logistic regression revealed that initial antibody levels (OR 1.39, 95% CI 1.09–1.76), graft function (OR 0.05, 95% CI 0.01–0.39), time after transplantation (OR 1.04, 95% CI 1.02–1.07) and MMF trough levels (OR 0.43, 95% CI 0.21–0.88) correlated with seroconversion, p<0.05. After controlling for these confounders, the effect of MMF dose reduction was calculated using propensity score matching. KTRs in the MMF reduction group had significantly lower MMF serum concentrations prior to the third vaccination and were more likely to develop antibody levels ≥35.2 BAU/ml than their matched KTRs (p=0.02).

**Conclusions:** Temporary reduction in MMF dose might be a promising approach to improve the immune response in KTRs.

## Introduction

Vaccination against SARS-CoV-2 (severe acute respiratory syndrome coronavirus type 2) is the most important and efficient strategy for preventing severe courses of COVID-19 infection (1–5). However, antibody levels decline within several months after vaccination (6–9) which is one of the main factors responsible for lower protection against novel virus variants (10). Susceptible patients’ groups such as kidney transplant recipients (KTRs) are less likely to develop protective immune response. Moreover, these patients demonstrate lower peak antibody values and faster antibody waning compared to the general population (7,11–13). To overcome this immunological deficit, intensified vaccination schedules with early booster vaccination are recommended. Despite this, seroconversion rates remain in a range between 25 to 50% (14–16), leaving still a large part of KTRs without adequate protection. One of the main factors interfering with the vaccine response is immunosuppressive treatment, especially with mycophenolate mofetil (MMF) and belatacept (11,17,18). In this prospective multicenter observational study, we aimed to determine the durability of immune responses to SARS-CoV-2 vaccine by measuring the kinetics of antibody waning among seropositive KTRs. Secondly, we assessed factors influencing the immune response to the third SARS-CoV-2 vaccination in previously negative KTRs. Finally, in a small group of KTRs, we analyzed whether the temporary reduction of MMF initiated by the treating nephrologist improved the immune response to the third SARS-CoV-2 vaccination.

## Methods

A total of 245 KTRs receiving two doses of either BNT162b2 (Biontech/Pfizer) or mRNA-1273 (Moderna) were included in this prospective multicenter observational study (NCT04743947). Results of the immune response to two vaccine doses have been published previously (11,19). KTRs included in the study had to be older than 18 years, without previous COVID-19 infection, and able to give informed consent for participation. All KTRs were on stable immunosuppressive medication and had no signs of acute graft rejection or a history of graft rejection within the last six months. The study was approved by the ethics committee of the Medical Faculty at the Heinrich-Heine University, Düsseldorf, Germany (study number 2020-1237) and in line with the Declaration of Helsinki, as revised in 2013.

### Seropositive KTRs after the first two SARS-CoV-2 vaccination

61 (24.9%) patients were seropositive after receiving two vaccine doses and 60 of them completed the follow up period. Two follow-up visits after the first vaccination were arranged, one after three months (average 94±8 days) and the other after six months (average 186±26 days) (Suppl. figure 1). At the follow-up visits, serum was collected for the measurement of humoral response. In addition, data on medication and laboratory parameters were derived from medical records (Suppl. figure 1).

### Seronegative KTRs after the first two SARS-CoV-2 vaccination

174 seronegative individuals after the two doses of vaccination received a third dose. 148 (85.1%) patients received a full dose of BNT162b2 (Biontech/Pfizer), 19 (10.9%) patients ChAdOx1 (AstraZeneca) and 7 (4%) patients Ad26.COV2.S (Johnson & Johnson). The time interval between the second and the third vaccine dose was 76±25 days. To assess the immune response and the kinetics of antibody waning, antibody levels were measured 22±7 days after the third vaccination and six months (185±10 days) after the first SARS-CoV-2 vaccination (Suppl. figure 1). At the visits data on medication and laboratory parameters were obtained from medical records. To evaluate factors associated with seroconversion induced by the third vaccination, a multivariate logistic regression model with backward elimination was performed. The following variables were included: age, gender, time after the transplantation, creatinine concentration, antibody concentration after second vaccine and variables associated with immunosuppression regimes such as trough MMF and tacrolimus concentrations in the serum, dual therapy as well as the use of calcineurin inhibitor, prednisolone or MMF therapy and their daily doses.

Due to the low immune response rate in KTRs following two SARS-CoV-2 vaccinations and growing evidence that the intensity of MMF therapy negatively affects humoral response rates, some attending nephrologists discussed with their patients the possibility to reduce temporarily the daily MMF dose and consulted the respective transplantation center. In selected cases and when approved by the patient recommendation to reduce the MMF dose three weeks prior to and untill one week after the third vaccination was given. According to this recommendation, we identified a group of 24 KTRs in whom the treating nephrologists have reduced the daily dose of MMF by 25-50%. In detail, there were 8 (33.3%) patients with 50% dose reduction, 9 (37.5%) patients with 33% dose reduction and 7 (29.2%) patients with 25% dose reduction. Based on the medical records, the immunosuppressive therapy was not significantly changed in any of the other KTRs. After controlling for baseline confounding factors such as antibody levels after the second vaccination, renal graft function, and MMF trough level (Table 2), we have estimated propensity scores for 174 patients who received the third SARS-CoV-2 vaccine dose. These scores were used to match 24 pairs with and without MMF reduction prior to the third vaccination. The representable matches were similar with respect to age, immunosuppression regime, MMF dose and trough levels before reduction, renal graft function, and initial antibody levels (Table 3).

### Evaluation of humoral response

All samples were tested for IgG antibodies against SARS-CoV-2 spike S1 subunit using Anti-SARS-CoV-2-QuantiVac-ELISA (Euroimmun AG, Lübeck, Germany) as well as for SARS-CoV-2 neutralization efficacy (NT) in a BSL-3 facility at the Institute of Virology, University Hospital Düsseldorf, Germany. Antibody levels above ≥35.2 BAU/ml were considered as positive which was in accordance with manufacturer’s instructions. Samples with antibody level >384 BAU/ml were further diluted 1:10 or 1:100 in IgG sample buffer. The lower detection limit was <3.2 BAU/ml. In addition to antibody level measurement, neutralizing capacity of the Anti-SARS-CoV-2 antibodies against was evaluated. An endpoint dilution neutralization test with the infectious SARS-CoV-2 B.1 (Wuhan Hu-1, GISAID accession ID: EPI_ISL_425126) and SARS-CoV-2 B1.617.2 (delta variant, GISAID accession ID:EPI_ISL_4471555) isolates at a TCID 50 of 100 was performed as described previously (20). The neutralization titer was determined as the highest serum dilution without virus-induced cytopathic effect (CPE).

### Data Analysis

Statistical analysis was performed using SPSS version 23 (SPSS Inc., Chicago, USA) and Graph Prism 9 (GraphPad Software, San Diego, USA). Shapiro–Wilk normality test was used to assess if data was normally distributed. Accordingly, continuous variables are expressed as mean ± standard deviation (SD) or median with interquartile range expressed as two numbers, Q1–Q3, respectively. Categorical variables are expressed as numbers (percentages). The difference between immune response groups was performed using Chi-square, unpaired t-test or Mann-Whitney test. The comparison of antibody levels and neutralizing capacity over the study period was performed using the Kruskal-Wallis test. Neutralizing antibody titers against Wuhan strain and delta variant at 6 months follow up visit were compared using Wilcoxon signed rank test. Multivariate logistic regression using backward elimination was used for indicating variables associated with a positive immune response after the third vaccine dose. These co-founding variables were further used for propensity matching process to assess the effect of MMF reduction on antibody development. The matching tolerance was set to 0.05. Receiver operating characteristic (ROC) curve of initial antibody levels and MMF trough concentration was used to predict seroconversion rate among KTRs. P values less than 0.05 were considered statistically significant.

## Results

The course of antibody levels within six months from the first vaccination was measured in the 60 KTRs who were seropositive after the second vaccination. This enabled the characterization of antibody waning (Figure 1A & B). After 3 and 6 months, antibody levels have decreased by 45% and 73%, respectively. Because of antibody waning 30% KTRs became seronegative over the follow-up period. In order to determine the antibody cut-off value in predicting the loss of serological status, analysis of ROC curve with area under the curve (AUC) was performed. AUC value was 0.763 (95% CI 0.639–0.887) whereas the optimal antibody cut-off value for becoming seronegative was ≤206.8 BAU/ml (Figure 1B) with the 83.3% sensitivity and 69% specificity. In line with this, the neutralizing capacity was diminishing (Figure 1C). Median neutralizing titers decreased from 1:20 (0–1:40) to 1:10 (0–1:40) in three months, p=0.7 and further to 0 (1-1:20) in six months, p<0.05. Moreover, the neutralizing capacity against the delta SARS-CoV-2 variant was significantly lower (p=0.016). Main factors associated with seroconversion to a formally negative status over the study period were initially low antibody levels as well as older age and impaired renal graft function. Interestingly, immunosuppression, especially the use of MMF was not associated with seroconversion (Suppl. table 1).

**Figure 1.**
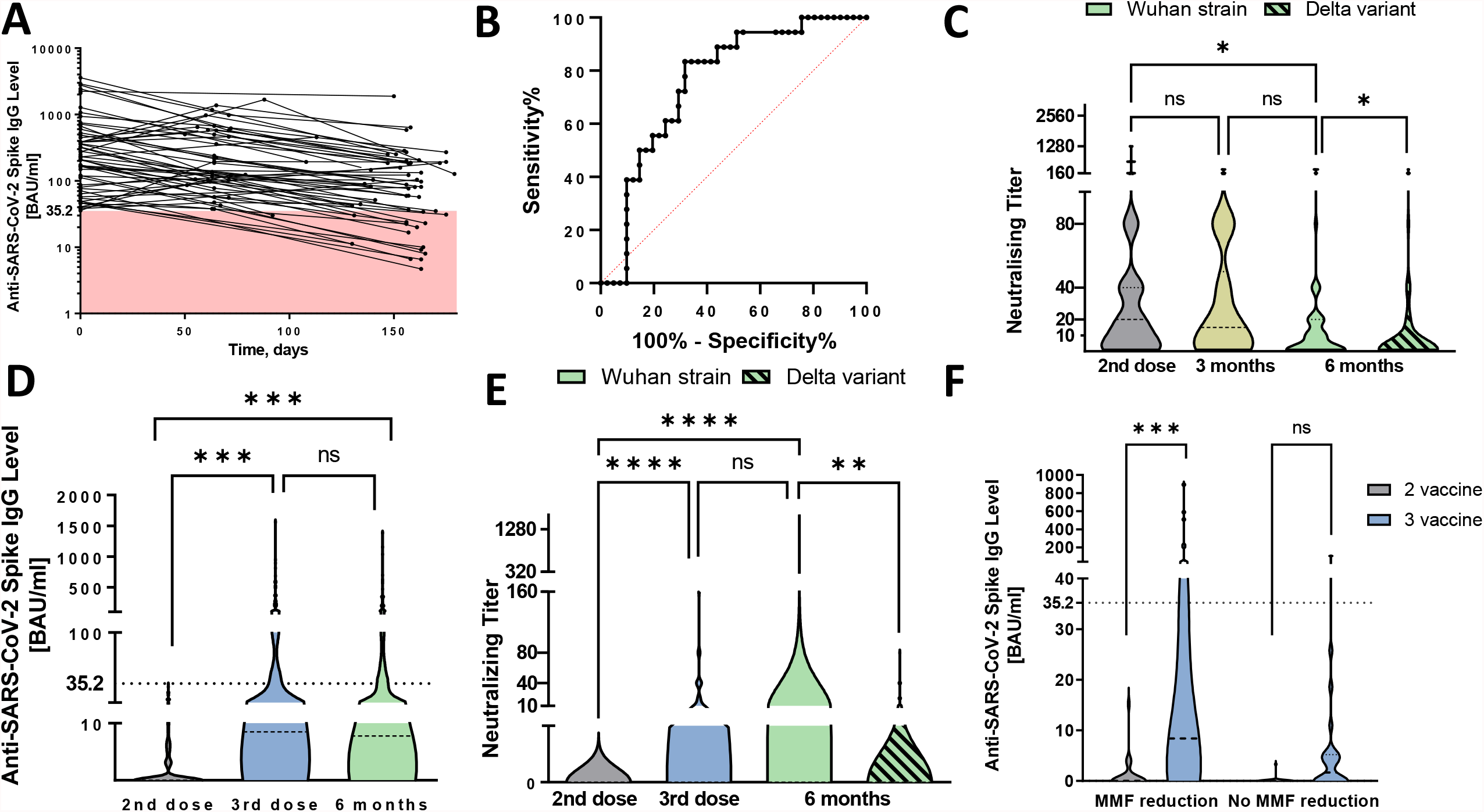
Antibody waning and humoral response to third SARS-CoV-2 vaccination among kidney transplant recipients. **A** Waning of anti-SARS-CoV-2 spike subunit S1 antibodies among 60 seropositive kidney transplant recipients (KTRs) after the first two vaccination over the follow-up period. The red colored space highlights antibody levels lower than the cut-off for seropositivity (<35.2 BAU/ml). **B** Receiver operating characteristics (ROC) curve for antibody level with respect to serological patient status at six month visit with AUC being 0.763 (95% CI 0.639–0.887), p=0.002. **C** Comparison of neutralizing antibodies among seropositive KTRs over the follow-up period. At the six months follow-up visit, serum was tested for neutralizing antibody capacity against the Wuhan strains and delta virus variant. **D** Comparison of antibody levels among KTRs after the third vaccination and at six months follow-up visit. Dashed line was set at 35.2 BAU/ml to outline seropositive patients. **E** Comparison of neutralizing titer among KTRs after the third vaccination and at six months follow-up visit. At the six months follow-up visit, patients’ serum was tested for neutralizing antibody capacity against the Wuhan strain and delta virus variant. **F** Comparison of antibody level between patients with and without MMF dose reduction. The graph represents immune response among 24 matched pairs using propensity score with respect to initial antibody concentration, renal graft function, time after transplantation and MMF trough level prior the dose reduction. Differences between different follow up visits were analyzed using Kruskal-Wallis test. Differences in neutralizing capacity against Wuhan strain and delta variant were assessed using Wilcoxon signed-rank test. **** represent p value < 0.0001, *** p < 0.001, ** p < 0.01, * p < 0.05.

174 previously seronegative KTRs received the third SARS-CoV-2 vaccination and have completed the follow-up period. According to the antibody levels detected 22±7 days post-vaccination, patients were divided into two groups: seronegative KTRs with antibody levels below 35.2 BAU/ml and seropositive KTRs with antibody level ≥35.2 BAU/ml. Baseline characteristics are presented in Table 1. In this group, 56 KTRs (32.1%) responded to the third SARS-CoV-2 vaccination with a median antibody level of 119 (76–353) BAU/ml and median neutralizing capacity of 1:10 (0–1:40) titer (Figure 1D & E). Antibody levels after the second vaccine and time after transplantation were the most relevant factors influencing seroconversion after the third vaccination (OR 1.39 (95% CI 1.09–1.76), p=0.006 and OR 1.04 (95% CI 1.02–1.07), p=0.001). Additionally, better renal graft function (OR 0.05, 95% CI 0.01–0.39), p=0.004 and lower trough MMF serum levels (OR 0.43, 95% CI 0.21– 0.88), p=0.021 were associated with higher antibody responses (Table 2). Daily MMF dose did not have a significant effect in the multivariate model, however in univariate regression analysis, daily MMF dose ≤1g showed 2.5 times higher odds for developing clinically relevant amount of antibodies, p=0.016.

**Table 1.** Characteristics of kidney transplant recipients according to immune response after the third SARS-CoV-2 vaccination. Patients were divided into 2 groups – the ones without any response to the vaccine or antibody level below 35.2 BAU/ml and patients with seroconversion. Seroconversion was defined as IgG antibody level against SARS-CoV-2 spike S1 subunit ≥35.2 BAU/ml. eGFR – estimated glomerular filtration rate, CNI – calcineurin inhibitor, mTOR - mammalian target of rapamycin, Aza – azathioprine, MMF – mycophenolate mofetil. Dichotomous data are presented as percentages whereas continuous data as means ± SD or median (Q1 – Q3). *** represent significant difference between the groups with p < 0.001, ** p < 0.01, * p < 0.05 using unpaired t – test, Chi-square test or Mann Whitney test.

| Immune response to third SARS-CoV-2 vaccination |  |  |
| --- | --- | --- |
| Parameter | < 35.2 BAU/ml | ≥ 35.2 BAU/ml |
| **Number of patients, N** | 118 | 56 |
| **Age, y** | 59.3 ± 13.1 | 59.8 ± 11.7 |
| **M:F** | 76:42 | 38:18 |
| **Time after transplantation, mo** | 46 (22 – 111) | 92 (56 -152)*** |
| **Creatinine, mg/dl** | 1.7 (1.3 – 2.2) | 1.4 (1.1 – 1.7)*** |
| **eGFR, ml/min/1.73m<sup>2</sup>** | 40 (29 – 50) | 54 (41 – 65)*** |
| **MMF concentration, µg/ml** | 2.7 (1.7 – 4.7) | 1.8 (1.4 – 2.5)*** |
| **Tacrolimus concentration, ng/ml** | 5.6 (5.0 – 6.5) | 5.6 (5.2 – 6.2) |
| **Antibodies after 2<sup>nd</sup> vaccination, BAU/ml** | 0 | 5.9 (0 – 15.2)*** |
| **Immunosuppression** |  |  |
| • **CNI** | 111 (94.1 %) | 56 (100 %) |
| • **Steroids** | 113 (95.8 %) | 51 (91.1 %) |
| • **mTOR/Aza** | 4 (3.4 %) | 2 (3.6 %) |
| • **MMF** | 111 (94.1 %) | 53 (94.6 %) |
| ≤ 1 g/d | 64 (57.7 %) | 41 (77.3 %)* |
| > 1 g/d | 47 (42.3 %) | 12 (22.7 %)* |
| • **Dual therapy** | 6 (2.5 %) | 6 (10.7 %) |
| With MMF | 4 (66.7 %) | 4 (66.7 %) |
| No MMF | 2 (33.3 %) | 2 (33.3 %) |
| • **Triple therapy** | 112 (94.9 %) | 50 (89.3 %) |
| With MMF | 107 (95.5 %) | 49 (98 %) |
| No MMF | 5 (4.5 %) | 1 (2 %) |

**Table 2.** Logistic regression model representing factors associated with the development of antibodies against SARS-CoV-2 spike S1 subunit after third vaccination among kidney transplant recipients. The model was performed using backward elimination process. Following variables have been included: age, gender, time after the transplantation, creatinine concentration, antibody concentration after second vaccination and variables associated with immunosuppression regime such as trough MMF and tacrolimus concentrations in the serum, dual therapy as well as the use of calcineurin inhibitor, prednisolone or MMF therapy and their daily doses. OR – odds ratio, CI – confidence interval. p values < 0.05 represent statistical significance.

| **Variables** | **$\beta$** | **OR** | **CI 95 %** | **p value** |
| --- | --- | --- | --- | --- |
| **Titer after 2<sup>nd</sup> vaccination** | 0.328 | 1.38 | 1.09 – 1.76 | **0.006** |
| **Cellcept concentration** | - 0.844 | 0.43 | 0.21 – 0.88 | **0.021** |
| **Creatinine concentration** | - 2.996 | 0.05 | 0.01 – 0.39 | **0.004** |
| **Time after transplantation** | 0.040 | 1.04 | 1.02 – 1.07 | **0.001** |

Next, we identified 24 KTRs who have been advised to reduce the daily MMF dose from three weeks prior until one week after the third vaccination (Table 3). The reduction in daily MMF dose resulted in a significantly decline in MMF serum concentrations (2.9 (1.7–4.0) μg/ml before reduction vs 2.1 (1.4–2.6) μg/ml after reduction, p<0.001). To analyze whether a temporary reduction in the daily MMF dose influences the immune response to the third SARS-CoV-2 vaccination, we used propensity score matching. After controlling for confounding factors (Table 2), we matched 24 pairs with and without MMF reduction. Baseline characteristics of matched KTRs are presented in Table 3. The group with MMF reduction showed significantly better responses compared to matched controls. Indeed, 7 (29.2%) patients achieved levels above the formal cut-off of a seropositive state after the daily MMF dose was reduced, whereas in the matched group only one patient (4.2%) developed antibody levels ≥35.2 BAU/ml (p=0.020, Figure 1F). In the MMF reduction group the 7 seropositive KTRs had lower MMF trough levels (1.6 (0.7-2.5) μg/ml) as compared to the 17 seronegative KTRs (2.3 (1.5-3.3) μg/ml, p =0.059). MMF trough levels of the 23 seronegative KTRs without MMF reduction were also significantly higher than those of the MMF reduction group (3.7 (2.3–5.4) μg/ml, p=0.016)

**Table 3.**
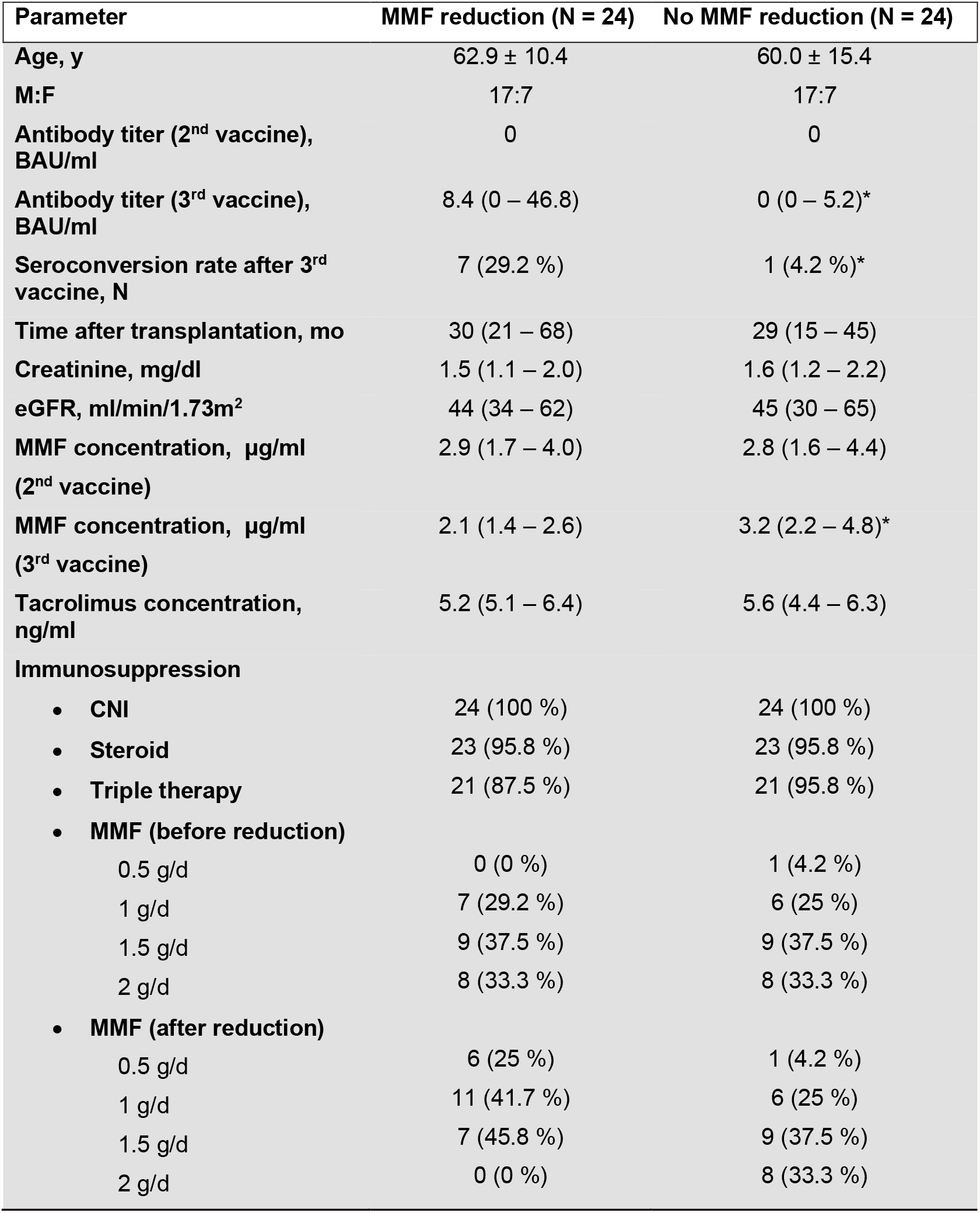
Characteristics of propensity matched groups with and without MMF dose reduction prior the third vaccination against SARS-CoV-2. Following variables have been included into matching process: creatinine concentration, MMF trough level, initial antibody level and time after the transplantation. Match tolerance was set to 0.05. Seroconversion was defined as IgG antibody level against SARS-CoV-2 spike S1 subunit ≥35.2 BAU/ml. eGFR – estimated glomerular filtration rate, MMF – mycophenolate mofetil, CNI – calcineurin inhibitor. Dichotomous data are presented as percentages whereas continuous data as means ± SD or median (Q1 – Q3). *** represent significant difference between the groups with p < 0.001, ** p < 0.01, * p < 0.05 using unpaired t – test, Chi-square test or Mann Whitney test.

| Parameter | MMF reduction (N = 24) | No MMF reduction (N = 24) |
| --- | --- | --- |
| Age, y | 62.9 ± 10.4 | 60.0 ± 15.4 |
| M:F | 17:7 | 17:7 |
| Antibody titer (2<sup>nd</sup> vaccine), BAU/ml | 0 | 0 |
| Antibody titer (3<sup>rd</sup> vaccine), BAU/ml | 8.4 (0 – 46.8) | 0 (0 – 5.2)* |
| Seroconversion rate after 3<sup>rd</sup> vaccine, N | 7 (29.2 %) | 1 (4.2 %)* |
| Time after transplantation, mo | 30 (21 – 68) | 29 (15 – 45) |
| Creatinine, mg/dl | 1.5 (1.1 – 2.0) | 1.6 (1.2 – 2.2) |
| eGFR, ml/min/1.73m<sup>2</sup> | 44 (34 – 62) | 45 (30 – 65) |
| MMF concentration, µg/ml (2<sup>nd</sup> vaccine) | 2.9 (1.7 – 4.0) | 2.8 (1.6 – 4.4) |
| MMF concentration, µg/ml (3<sup>rd</sup> vaccine) | 2.1 (1.4 – 2.6) | 3.2 (2.2 – 4.8)* |
| Tacrolimus concentration, ng/ml | 5.2 (5.1 – 6.4) | 5.6 (4.4 – 6.3) |
| **Immunosuppression** |  |  |
| • CNI | 24 (100 %) | 24 (100 %) |
| • Steroid | 23 (95.8 %) | 23 (95.8 %) |
| • Triple therapy | 21 (87.5 %) | 21 (95.8 %) |
| • MMF (before reduction) |  |  |
| 0.5 g/d | 0 (0 %) | 1 (4.2 %) |
| 1 g/d | 7 (29.2 %) | 6 (25 %) |
| 1.5 g/d | 9 (37.5 %) | 9 (37.5 %) |
| 2 g/d | 8 (33.3 %) | 8 (33.3 %) |
| • MMF (after reduction) |  |  |
| 0.5 g/d | 6 (25 %) | 1 (4.2 %) |
| 1 g/d | 11 (41.7 %) | 6 (25 %) |
| 1.5 g/d | 7 (45.8 %) | 9 (37.5 %) |
| 2 g/d | 0 (0 %) | 8 (33.3 %) |

## Discussion

First, we investigated the decline in antibody titers among KTRs over time and determined factors predicting this decline and seroreversion, respectively. Results from the general population showed that antibody levels decline significantly within six months after the second vaccination (7,21). In our cohort, 30% KTRs seroconverted to a negative state within six months from the first vaccination. The most important factor for predicting the decline below the threshold of 35.2 BAU/ml was the initial antibody level. An antibody level ≤206.8 BAU/ml predicted seroreversion with 83.3% sensitivity and 69% specificity. In addition, older KTRs and KTRs with impaired graft function seroreverted more frequently. Notably, the kinetic of antibody waning in KTRs of the present study is similar to the one reported in the general population (22,23). However, since KTRs develop lower antibody levels, they are more predisposed to seroreversion (11). Therefore, shorter intervals of booster vaccinations should be recommended for KTRs. Measurements of the neutralizing capacity showed similar dynamics. Neutralizing antibody levels, predictive for the immune response in symptomatic SARS-CoV-2 infection (24), decline rapidly in KTRs. Of note, we could also show that the neutralizing capacity in KTRs depends on the virus variant. Patients had a lower amount of neutralizing antibodies against the delta SARS-CoV-2 variant which is in accordance with previous studies (10,25).

Second, we analyzed the immune response to a third SARS-CoV-2 vaccination among 174 seronegative KTRs. 32.1% of KTRs seroconverted after the third vaccination which is almost the same rate as observed after the second vaccination (11,12,26). These results are in line with previously published studies representing smaller KTRs cohorts (14–16,27). Despite the longer interval between 2nd and 3rd vaccination in the present study, the mean antibody levels were comparable to results from studies in which the third vaccination was administered earlier (16,27). Therefore, other factors seem to influence the antibody development. The lack of immune response to the first two SARS-CoV-2 vaccinations in KTRs is related to the use of immunosuppressive drugs, especially MMF (11,13,28). We observed a similar association for the third vaccination. An increase in trough MMF concentrations by 1μg/ml was associated with an almost 60% lower seroconversion. Additionally, already detectable antibodies below the formal cut-off (between 3.2 and 35.2 BAU/ml) prior to the third vaccination were also associated with a better humoral immune response. Better renal graft function and longer time after transplantation were also related to success of the third vaccination, similar to results observed after two vaccinations (11,17).

The importance of MMF affecting the vaccination response in KTRs was further analyzed in 24 KTRs in whom the attending nephrologists had temporarily reduced the MMF dose. Since this group was rather small, we used propensity score matching to address the effect of MMF reduction. As expected, KTRs of the MMF reduction group had significantly lower MMF serum concentrations prior to the third vaccination than patients without reduction. Importantly, these patients were more likely to develop antibody levels ≥35.2 BAU/ml. Seroconverting patients had at least a 33% dose reduction and average MMF through levels < 2 μg/ml suggesting that a minimum MMF reduction may be required for achieving an immune response. As such low MMF concentrations might bear a risk for graft rejection, close monitoring of graft function is necessary (29). In a recent study, the effect of MMF was addressed by pausing the treatment one week prior untill one month after the fourth vaccine dose. Using this approach, seroconversion rate among KTRs increased from 34.5 % on day seven to 76 % at the one month follow-up visit (30) suggesting that MMF pause for more than one week may further improve seroconversion rate in KTRs. Nevertheless, larger randomized controlled studies are necessary and already planned to address this association in more detail (31).

There are some limitations of the present study. We have performed two follow-up visits. More frequent visits may resolve antibody waning more precisely. In addition, we cannot exclude a bias in the process of MMF reduction. Each decision was made by the treating nephrologist after contacting the transplantation center considering each case individually. Another relevant point is that our MMF reduction group was relatively small with different categories of dose reduction. Finally, we only studied humoral immune responses and it is unclear if cellular immunity against SARS-CoV-2 follows the same pattern in KTRs.

In summary, we showed that KTRs have antibody kinetics similar to the general population. However, these patients develop lower antibody levels, which is the main reason for shorter humoral protection. In addition, the third vaccination against SARS-CoV-2 leads to seroconversion in 32.1% in previously seronegative patients. This is a substantial achievement, however many KTRs are still insufficiently protected. Our results suggest that a moderate, temporary MMF dose reduction could be a safe approach to improve vaccination success in KTRs.

## Data Availability

The data that support the findings of this study are available at: doi:10.17632/g3wpr3xtpx.1

## Author contributions

Conceptualization: TK, KI, HMO, OA, LCR, JT, JS Formal analysis: MK, LM, JL, SF Investigation: MK, LM, JH, JL, SF, TK, KI, NL, MK, MS, CS, TF, MA, HMO, EK, MK, OA, JT, JS Validation: LM, HS, JT Writing – original draft preparation: MK, LCR, JT, JS Writing - review and editing: MK, LM, JH, JL, TK, KI, MK, HS, NL, MS, CS, TF, MA, HMO, OA, TL, EK, MK, LCR, JT, JS Resources: TL Supervision: JT, JS

## Acknowledgment

The authors kindly thank for the employees of nephrology ambulance of University clinics of Duesseldorf, KfH and Nephrocare Dialysis centers as well as employees of Solingen hospital for their support with organizing patients’ visits. We are thankful for Yvonne Dickschen for her technical assistance.

## Data sharing statement

Raw data were generated at University Hospital Düsseldorf, Heinrich-Heine-University Düsseldorf, Düsseldorf, Germany. The data that support the findings of this study are available at: doi:10.17632/g3wpr3xtpx.1

## Supplemental Table of Contents

**Supplemental Table 1.** Characteristics of 60 seropositive kidney transplant recipients at 6 month follow up visit with respect to serological status. Seropositivity was defined as IgG antibody against SARS-CoV-2 spike S1 subunit titer above 35.2 BAU/ml. eGFR – estimated glomerular filtration rate, Δ change during follow up period, CNI – calcineurin inhibitor, mTOR – mammalian target of rapamycin, Aza – azathioprine, MMF – mycophenolate mofetil. Dichotomous data are presented as percentages whereas continuous data as means ± SD or median (Q1 – Q3). *** represent significant difference between the groups with p < 0.001, ** p < 0.01, * p < 0.05 using t – test, Chi-square test or Mann Whitney test.

| Parameter | Seropositive (N = 42) | Seronegative (N = 18) |
| --- | --- | --- |
| Age, y | 57.5 ± 12.5 | 67.2 ± 8.0 ** |
| M:F | 28:14 | 11:7 |
| Antibody titer, BAU/ml | 337 (141 – 630) | 94 (49 – 181) * |
| Antibody titer at 6 months, BAU/ml | 152 (82 – 251) | 15 (8 – 25) *** |
| Neutralizing antibodies at 6 months | 1:10 (0 – 1:20) | 0 |
| Neutralizing antibodies against Delta variant at 6 months | 1:10 (0 – 1:20) | 0 |
| Time after transplantation, mo | 154 (91 – 234) | 187 (131 – 266) |
| Creatinine, mg/dl | 1.4 (1.1 – 1.7) | 1.7 (1.5 – 2.2) * |
| eGFR, ml/min/1.73m<sup>2</sup> | 48 (39 – 60) | 36 (29 – 44) * |
| Δ Creatinine, mg/dl | 0.06 ± 0.25 | -0.01 ± 0.35 |
| **Immunosuppression** |  |  |
| • CNI | 40 (95 %) | 18 (100 %) |
| • Steroids | 38 (90 %) | 18 (100 %) |
| • mTOR/Aza | 2 (4.7 %) | 3 (16.7 %) |
| • MMF | 25 (59.5 %) | 4 (22.2 %) ** |
| ≤ 1 g/d | 23 (92 %) | 4 (100 %) |
| > 1 g/d | 2 (8 %) | 0 (0 %) |
| • Dual therapy | 19 (45.2 %) | 11 (61.1 %) |
| With MMF | 3 (15.8 %) | 0 (0 %) |
| No MMF | 16 (84.2 %) | 11 (100 %) |
| • Triple therapy | 23 (54.8 %) | 6 (33.3 %)* |
| With MMF | 22 (95.6 %) | 4 (66.7 %) |
| No MMF | 1 (4.4 %) | 2 (33.3 %) |
| • Monotherapy | 0 (0 %) | 1 (5.6 %) |

**Supplemental Figure 1.**
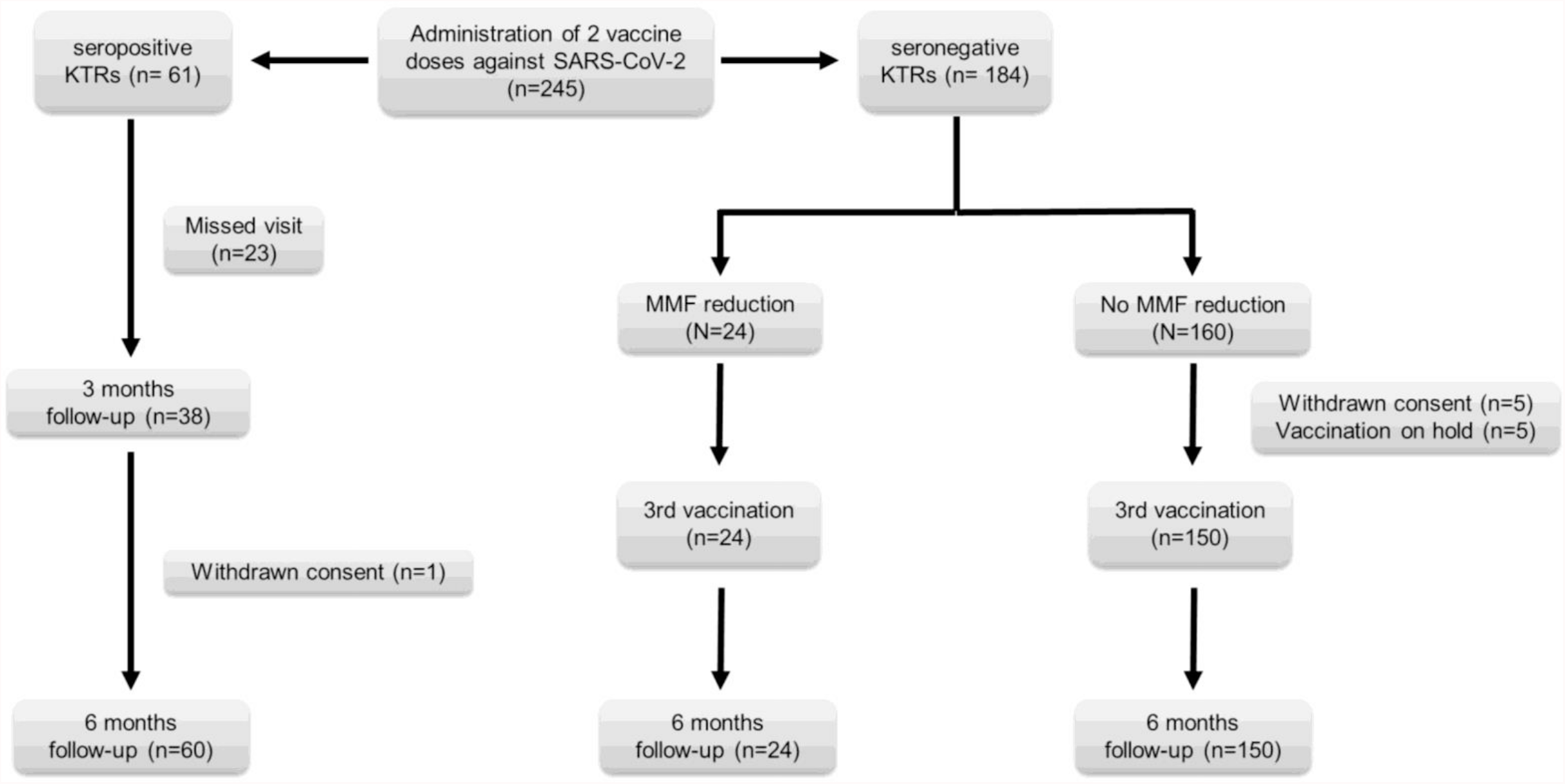
Flow chart demonstrating study design.

